# Impulse dispersion of aerosols during playing wind instruments

**DOI:** 10.1101/2021.01.25.20248984

**Authors:** Sophia Gantner, Matthias Echternach, Reinhard Veltrup, Caroline Westphalen, Marie Christine Köberlein, Liudmila Kuranova, Gregor Peters, Bernhard Jakubaß, Tobias Benthaus, Michael Döllinger, Stefan Kniesburges

## Abstract

Musical activities especially singing and playing wind instruments have been singled out as potentially high-risk activities for transmission of SARS CoV-2, because of a higher rate of aerosol production and emission. Playing wind instruments can produce condensation water, droplets of saliva, and aerosol particles, which hover and convectional spread in the environmental air and can be potentially infectious.

The aim of this study is to investigate the primary impulse dispersion of aerosols during playing different wind instruments in comparison to breathing and speaking. Nine professional musicians (3 trumpeters, 3 cross flutists and 3 clarinetists) of the Bavarian Symphony Orchestra performed the main theme of Ludwig van Beethoven‘s 9^th^ symphony, 4^th^ movement in different pitches and loudness. Thereby, the inhaled air volume was marked with small aerosol particles produced with a commercial e-cigarette. The expelled aerosol cloud was recorded by cameras from different perspectives. Afterwards, the dimensions and dynamics of the aerosol cloud was measured by segmenting the video footage at every time point.

Overall, the cross flutes produced the largest dispersion at the end of task of up to maximum distances of 1.88 m in front direction. Thereby it was observed an expulsion of aerosol in different directions: upwards and downwards at the mouthpiece, at the end of the instrument and along the cross flute at the key plane. In comparison, the maximum impulse dispersion generated by the trumpets and clarinets were lower in frontal and lateral direction (1.2 m and 1.0 m in front-direction). The expulsion to the sides was also lower. Consequently, a distance of 3 m to the front and to the sides of 2 m for the cross flutes in an orchestral formation is proposed, for trumpets and clarinets a safety distance of 2 m to the front and 1.5 m between instrumentalists are recommendable.

## Introduction

In order to limit the person-to-person transmission of COVID-19, gatherings of people, like it is common in cultural institutions and music events, have been restricted worldwide. Transmissions of COVID-19 are suspected to happen by direct contact, by small saliva droplets with diameters ≥5 μm and aerosols with diameters ≤ 5 μm [1]. As some outbreaks of the disease were associated with rehearsals of choirs or concerts [2, 3, 4], these events have been singled out as potentially high-risk activities for transmission of SARS CoV-2, which was confirmed by the fact that singing is associated with a much higher rate of aerosol emission than breathing and speaking [5, 6].

Similarly, playing wind instruments can produce aerosols, droplets of saliva and condensation water, which can be potentially infectious if the musician is infected with the SARS CoV-2virus, even without symptoms. Whereas the droplets in general fall to the ground after a certain distance, the aerosols remain hovering in the air owing to their small mass and convectively spread in the environment. Consequently, infections appear possible due to exposure or inhalation of virus containing aerosols in closed rooms.

Particularly, straight, long instruments with conically increasing diameter like vuvuzelas have the capacity to propel very large numbers of aerosols into the air which are able to penetrate the lower lung of another person [7]. In contrast, a few studies measuring the concentration and the size of airborne particles from brass and woodwind instruments reported low emission rates at distances of .5 m to 4 m [8]. Ruichen et al. categorized 15 different wind instruments into low (tuba), intermediate (bassoon, piccolo, cross flute, bass clarinet, french horn and clarinet) and high risk (trumpet, bass trombone and oboe) levels based on a comparison of their aerosol generation with those from normal breathing and speaking [9].

Previous studies measured air velocity and movement emerging from wind instruments using photography, Schlieren optics, sensors in the vicinity of the musician and Particle Image Velocimetry [10, 11, 12, 13, 14]. The latter observed airflow up to .5 m from the bell of brass instruments like trumpet, trombone and euphonium. In woodwinds, especially the cross flute caused an airflow over a distance of more than 1 m [13]. In a previous study [15], artificially added aerosols were analyzed with respect to the impulse dispersion characteristics in professional singers for singing and speaking tasks. It was found that the maximum impulse dispersion to the front was up to 1.4 m for singing and less pronounced to the sides. For wind instruments, however, the impulse spreading characteristics of aerosols have not yet been investigated. The aim of this study is to reveal the aerosol expulsion characteristics of wind instruments. The intention was to show the spreading of the expelled potentially virus-laden aerosol particles immediately during playing. Based on this knowledge, safety distances can be defined for other members of an orchestra to prevent them to stay in the primary aerosol cloud and therewith to reduce the infection risk with SARS-CoV-2.

## Material and Methods

With the approvement from the local ethical committee (20-395), 9 professional musicians (1 female, 8 male, age 48±4 years) from the Bavarian Symphony Orchestra (Symphonieorchester des Bayerischen Rundfunks) were included in the study. All of them were non-smokers without pulmonic symptoms. Three of them were trumpeters, three cross flutists and three clarinetists. All participants practice western classical music and had completed their instrumental studies at an University of Music. None of them suffered from dysphonia at the time of the study: The values for the Voice Handicap Index 12 (VHI-12) in a German version [16] exhibited norm values (mean 3 SD4).

### Tasks

All participants performed a part of the main theme (melody, M) of the fourth movement of Ludwig van Beethoven‘s 9^th^ symphony in D major key. The musicians played the melody in their specific ranges of “high” (h) and “low” pitch (l). Additionally, they were asked to play the high pitch task in a loud (Mh+) and a soft version (Mh-). Summarizing up there were three different tasks for each instrument: melody on a high pith and loud (Mh+), melody on a high pith and soft (Mh-) and melody on a low pitch and loud (Ml+).

The specific ranges for each instrument are as follows: (Mh: trumpet: absolute pitch range: D5-A5, fundamental frequency *f*_o_ range: 587-880 Hz; Mh: cross flute: D6-A6, 1175 – 1750 Hz; Mh: clarinet: Bb4-F5, 466 - 699 Hz); (Ml: trumpet: D4-A4, 294 - 440 Hz; Ml: cross flute: D5-A5, 587 - 880 Hz; Ml: clarinet: Bb3-F4, 233 - 349 Hz).

In addition, there were specific tasks for each instrument:

- The trumpeters performed two further tasks, by playing with the mouthpiece only and without the mouthpiece by just buzzing the melody with the lips.
- The clarinetists played the melody with mouthpiece only as the trumpeters. Additionally, they produced a low E, where all keys are closed
- The cross flutists performed the task with the so called fluttering tongue technique.

All these tasks lasted approx. 6 s corresponding to a speed of 80-90 bpm for the M tasks.

Beside the instrumental tasks, each musician spoke the affiliated original text (T) („Freude, schöner Götterfunken, Tochter aus Elysium“) of Friedrich Schillers poem “An die Freude” on comfortable pitch and average loudness.

In a pre-experiment the Forced Expiratory Pressure in 1 Second (FEV1) and Tiffeneau-Index (TIFF Index= FEV1/VC) were measured in order to investigate the pulmonary function with a spirometer (ZAN100, Oberthulba, Germany).

### Test setup

For the experiments, all subjects inhaled the smoke of an e-cigarette filled only with the basic liquid, which consists of 50 % glycerin and 50 % propylene glycol. A Lynden Vox e-cigarette was used to nebulize this liquid, (Lynden GmbH, Berlin, Germany). The particles generated in e-cigarettes have a diameter in the range of 250 - 450 nm and are thus in the range of aerosols expelled during breathing and talking [15, 17]. For each task a spirometer (ZAN100, Oberthulba, Germany) was coupled to the mouthpiece of the e-cigarette to measure the individual inhaling volume. Each musician stood on a lifting platform to lift them to the correct height. On the height of the forehead, a mark was mounted to show the musician’s correct position on the platform, as shown in the top view part of Fig. 1. While standing on the platform, each musician inhaled a similar volume of smoke with an average of 0.76 litres (SD 0.26 litres) of smoke controlled by the spirometer. Immediately after inhalation, each musician moved to the mark and started to perform the task. After they had finalized the task, the musicians were instructed to maintain their position on the platform without moving for 60 s with the aim to trace the exhaled cloud. Because all musicians were non-smokers, they were instructed before the actual experiments how to inhale correctly. In cases of sudden coughing, the task was repeated until it was performed correctly.

**Figure 1:**
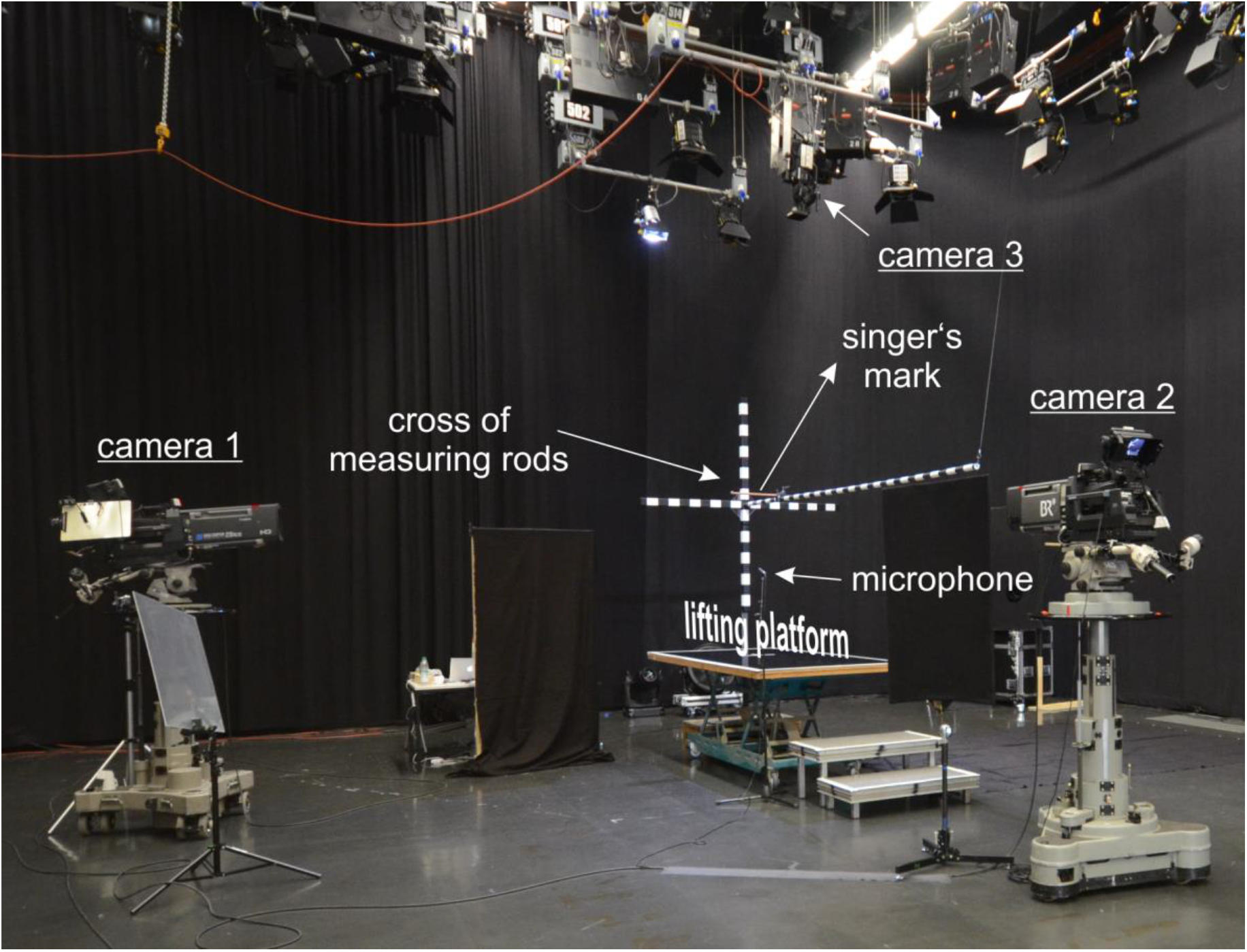
Picture of the test setup including the three cameras, the lifting platform and measuring rod cross (spatial directions are indicated by x, y and z).

All measurements were conducted in a studio with the dimension of approximately 27 m x 22 m x 9 m (width x length x height). The setup is depicted in Fig. 1. The distance to the wall behind the platform was 4 m and 5 m to the left. The distance between the platform and the frontal and the right wall was more than 7 m. Three measuring rods for each spatial direction were mounted at the lifting platform to convert the pixel dimension of the recorded picture into metric dimensions. The rods were assembled in a cross configuration and had a 2 cm and 10 cm scale, respectively.

Three full HD Sony HDC 1700R cameras of the Bavarian Broadcast (resolution 1920 x 1080 pixels) recorded the musicians and the cloud from a side, a top and a front view perspective as shown in Fig. 1. The side and front cameras were equipped with Canon DIGI SUPER 25 XS lenses (Canon, Tokio, Japan) and the top camera with a HD Fujinon HA14×4.5BERM/BERD wide angle HD lens (Fujinon, Tokio, Japan). All cameras recorded synchronously at a frame rate of 25 fps.

The audio signal was recorded with a Sennheiser KMR 81 directional and a ME62 omnidirectional microphone (Sennheiser electronic GmbH, Wedemark, Germany) which was placed at the lower right corner of the lifting platform, in a distance of approx. 1.5 m to the musician’s mouth, as shown in the top view zoom in Fig. 1. For estimation of the sound pressure level (SPL), the microphones were calibrated with the Sopran software (Svante Grandqvist, Karolinska Institut, Stockholm, Sweden) using a sound level meter (Voltcraft, Hong Kong, China).

In order to achieve a high contrast between the cloud of smoke and the background, the entire studio was covered with black linen and the musicians wore black clothes. The smoke was illuminated with three spotlights positioned on the left hand side of the platform: one in the left corner behind the platform with a distance of approx. 5 m to the musician: one in front of the platform on the left hand side in a distance of 7 m and another directly behind the platform approx. 3.5 m from the rear edge of the platform.

Before each task, the studio was aerated with the main gate open for at least 2 min using a fan behind the platform. The main gate was at the opposite side of the studio with a distance of approx. 20 m to the platform. Afterwards, the gate was closed and all participants were instructed to stop moving for another 2 min to let air circulation settle down. Subsequently, the task started with the musician’s inhalation of the e-cigarette basic liquid. The temperature and the humidity in the studio were measured. The mean temperature was 23 °C (SD .46) and the relative humidity 46 % (SD .95).

### Segmentation analysis

The expelled vapor cloud was segmented in footage of the side and top view cameras by an in-house software tool, yielding its contour as a function of time in order to track its temporal evolution. Therefore, (1) the side and top camera recordings were converted into gray scales and (2) the subjects and bright equipment parts in the region of interest (i.e. parts of the subjects’ skin of their hands, the white sectors of the measuring rods) were covered with black masks using the software Sensarea (Grenoble Institute of Technology (INPG), France). A coordinate system was defined with its origin at the subjects’ mouth. Then, the cloud contour was segmented in each frame of a video using a threshold-based region-growing algorithm.

After the segmentation, the maximum expansions of the cloud in each frame were identified in all three spatial directions (x, y, z). In the final results showing the maximum cloud expansion from the mouth several outlieres occurred if non-smoke regions were segmented when the cloud reached or non-cloud regions with similar grayscale value. Those outliers were removed in two steps: (1) a moving median filter with a fixed window length of 30 time points and (2) a subsequent cubic spline fitting approximation. The computation of curve smoothing was performed in Matlab (The Mathworks Inc., Natick, MA).

## Results

In the pre-experiment (spirometry only) without inhalation of the basic e-cigarette liquid, the subjects showed lung function parameters with a median FVCin of 4.53 litres (SD= 0.66 litres) and a TIFF Index of 86 (SD 4.3). The volume of basic liquid inhaled with an e-cigarette immediately before each task was on average 0.76 litres (SD 0.26 litres).

All subjects succeeded the tasks without major problems, even though the sound generation properties of the instruments changed by the higher density of the smoke in comparison to air. For the breathing task, a mean distance over all subjects of 1.26 m (SD .51 m) in the front (x), .41 m (SD .24 m) in vertical (z) and .33 m (SD .32 m) in horizontal, transversal direction (y) were detected. The mean distance for speaking amounts .59 m (SD .24 m) to the front (x), .43 m (SD .17 m) in vertical (z) and, .29 m (SD .10 m) in horizontal transversal direction (y). These data were used for comparison with the following measurements during instrumental playing.

For the evaluation of the playing tasks Mh+, Mh- and Ml+, the distances were taken at the end of the task which represents the time point 0 in Fig. 2 - 4.

**Figure 2:**
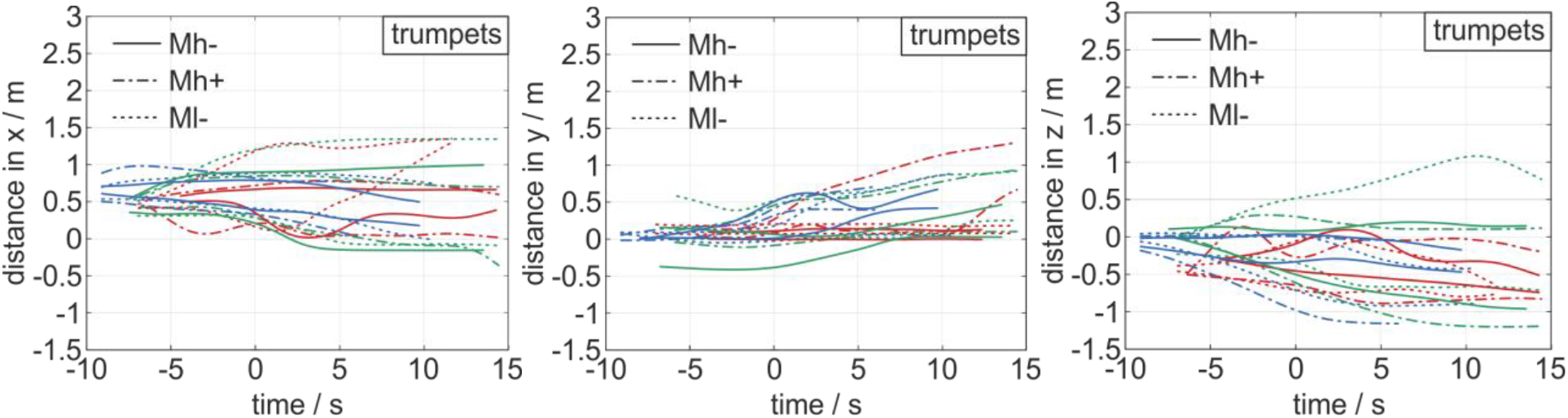
Diagrams of distances in x-, y- and z-direction from left to right for the trumpets. The 0 point in the time-scale refers to the end of the task. Each task is represented by two lines showing the lower and upper border of the cloud in the respective direction. The solid line represents task Mh-, the dashed line task Mh+ and the dotted line task Ml+. The different colors indicate the different players.

### Trumpets

Figure 1 shows the representative images in different camera views while the Mh+ task is performed by a male subject (the corresponding videos can be found in the supplemental data).

The median distances of the aerosol impulse dispersion generated from the three trumpeters during the playing tasks Mh+, Mh- and Ml+ amount .86 m in x-, .30 m in y- and .02 m in z-direction at the end of the task. The maximum distances were 1.2 m in x-, .52 m in y- and .52 m in z-direction.

The time evolution of the aerosol cloud is shown in Fig. 2 for each player and the tasks Mh+, Mh- and Ml+. In all spatial directions, the main distance was generated before the end of task at time point t=0 s. In the further progress, the motion of the cloud decelerated and reached a constant distance at t=15 s. In some cases, the distance becomes smaller after the task end which is caused by the spreading of the aerosol particles and the resulting thinning of the cloud. In those cases the cloud vanished and could not be segmented any more.

The curves representing the distance in x, start at x=.5 m, Fig. 2 a). The reason is that the origin of the coordinate system is placed at the player’s mouth, but the cloud predominantly exited at outlet of the trumpet approx. 50 cm further downstream in x-direction.

In the y-direction, the cloud tended to move in positive direction even after the task has ended. This tendency can also be seen in Fig. 3 b) and 4 b) for the cross flutes and clarinets. In accordance to the study with the singers [15], it could be assumed that this cloud motion was potentially produced by the motion of the player from the rear end of the platform (where he/she inhaled the aerosols) to the mark.

**Figure 3:**
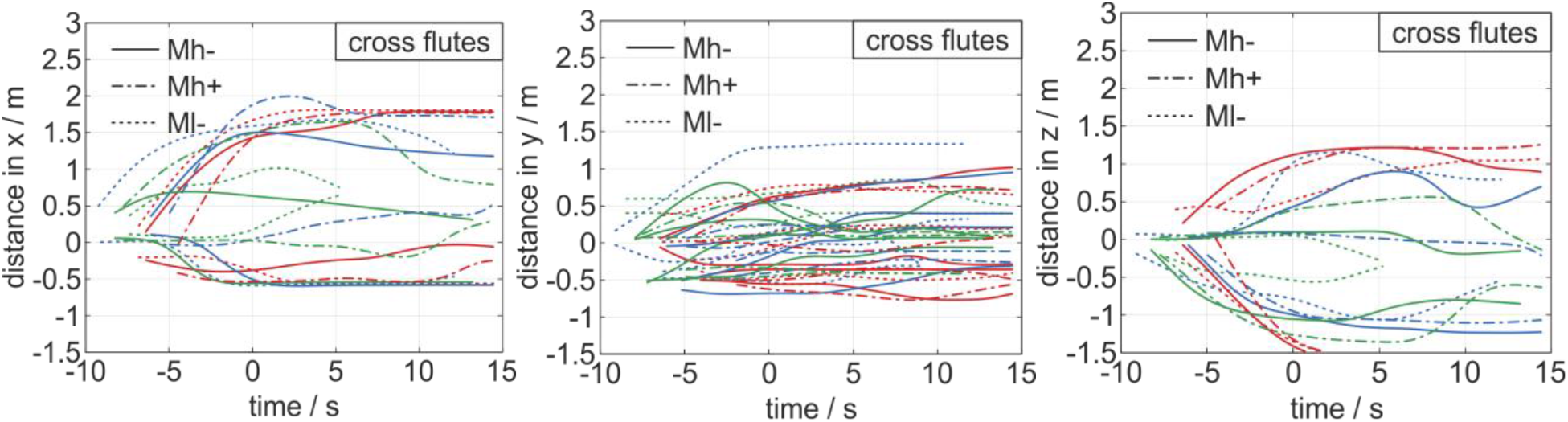
Diagrams of distances in x-, y- and z-direction from left to right for the cross flute. The 0 point in the time-scale refers to the end of the task. Each task is represented by two lines showing the lower and upper border of the cloud in the respective direction. The solid line represents task Mh+, the dashed line task Ml+ and the dotted line task Mh-. The different colors indicate the different players.

Regarding the vertical direction, the cloud tended to move in positive z-direction. The tasks of playing with the mouthpiece only and with just buzzing achieved similar distance of .88 m and .84 m in x-direction, respectively. The expansion in y-direction was found to be slightly larger with .5 m and .47 m. The expansion in z-direction was .06 m and .03 m.

### Cross flutes

In the case of the cross flutes, an expulsion of aerosol in different directions has been observed: upwards and downwards at the mouthpiece, at the end of the instrument and along the flute at the key plane. Overall, the flutes produced the largest median distance at the end of task of 1.51 m in the front (x), .64 m in lateral direction (y) and .02 m in vertical direction (z) considering all three playing tasks and players. The maximum distance was 1.88 m in x-, 1.29 m in y- and 1.12 m in z-direction (Figure 3).

The large distances of the aerosol motion in x-direction was achieved by the expulsion at the mouthpiece owing to the high velocity of the player’s expiration flow. The aerosol ejection at the key plane and the end of the flute occurred with a much smaller velocity.

Regarding the temporal evolution of the aerosol cloud spreading, the main distance is again generated in the period until the end of task (t=0 s). Afterwards the cloud decelerated to reach a constant distance or even dissolved represented by a decrease in distance. The motion trend in positive y-direction can also be seen but is not as distinct as for the trumpets and clarinets.

As an additional experiment the flutists played in flutter-tongue style. This resulted in similar median distances of 1.56 m in x-, .40 m in y- and .56 m in z-direction compared with the normal playing style.

### Clarinets

The clarinets showed aerosol ejections at the mouthpiece, at the end of the instrument (main outlet) and at the key plane. The median distances of all tasks and players were measured to be 1.51 m in x-, .55 m in y- and .51 m in z-direction. The maximum distance in x-direction reached by a clarinet player was 1.0 m, 0.95 m in y- and 1.1 m in z-direction.

Figure 4 shows the temporal evolution of the aerosol expansion during playing the clarinets: The main distance is reached before the end of task with a subsequent deceleration resulting in a constant distance or even decay of the cloud resulting in a decrease of distance, predominatly for the x-direction. The motion tendency in positive y-direction is again clearly visible, as shown in Fig. 4 b).

**Figure 4:**
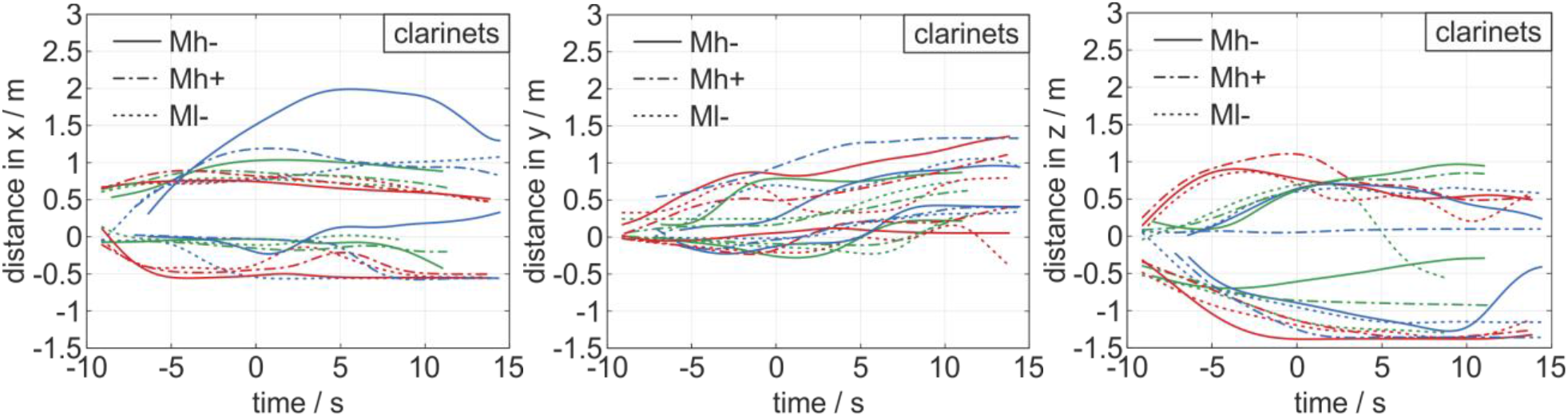
Diagrams of distances in x-, y- and z-direction from left to right for the clarinet. The 0 point in the time-scale refers to the end of the task. Each task is represented by two lines showing the lower and upper border of the cloud in the respective direction. The solid line represents task Mh+, the dashed line task Ml+ and the dotted line task Mh-. The different colors indicate the different players.

For the additional tasks of playing with mouthpiece only, a median distance of .90 m in x-, .86 m in y- and .80 m in z-direction was observed. Furthermore, the cloud was examined for playing a low E, with all keys kept closed. As result, the entire air flow exited via the main outlet at the end of the instrument. Here, similar values, compared to the main tasks with median distances of .96 m (x), and .17 m (y) were detected.

## Discussion

Musical activities especially singing and playing instruments, are assumed to contribute to the transmission of CoVID19 by aerosols [18-20]. The presented data showed that the expulsion and spreading of aerosols during playing the cross flute reached for individual cases, in a short amount of time, distances to the front of up to 1.88 m, for trumpets up to 1.2 m and for clarinets up to 1.2 m. For one clarinet player, the cloud spread up to 1.51 m as shown in Fig. 2 a, but this can be explained by convection flow to the left and front as explained below. Furthermore, the data reveal that the distance of dispersion is similar for loud and soft playing as well as playing only with mouthpiece. In contrast speaking showed much lower dispersions.

In order to protect other people from potentially infectious particles during the CoVID19 pandemic, it seems important to understand the impulse dispersion dynamics for aerosols since the aerosol density is mostly highest in the primary expelled cloud. Consequently, the distances reported here could be used in order to define the near field with the highest aerosol density and to derive recommendations concerning safety distances. The presented data showed that for all instruments the impulse dispersion to the front was below 1.5 m at the end of the task. The only exception was the cross flute reaching distances up to 1.88 m. While a safety distance for the clarinets and trumpets could be recommended by 2 m, the distance for the cross flute should be chosen greater with a distance to the front of 3 m. The lateral distances to players’ sides were found much lower. In this respect, at least 1.5 m to these directions should be defined to exclude a presence of a neighbor player in the near field. Because the cross flute produced three different clouds, the distance to the side should be estimated greater with 2 m. The aerosol dispersion reached down to the floor and 1.2 m above the instrument. Because usually the person-to-person distance in the vertical direction plays a relatively minor role, the recommendations for safety distances could be considered minor relevant for safety distances. However, it should be taken into account that the flutists are not positioned additionally elevated in the back rows of the orchestra formation, since the main direction of flow of the aerosols is directed forward and downward at an angle of about 30°-50°.

The transmission during the pandemic is not only dependent on the impulse characteristics analyzed in the presented study but also on the absolute aerosol generation during playing. He et al. [9] measured the absolute aerosol generation and estimated that the trumpet (among bass trombone, and oboe) is one of the high risk instruments. The aerosol production has been assumed to be caused by the lip oscillation during trumpet playing. However, in the presented study the largest amount of the aerosol seemed to remain in the trumpet and only a very small number of aerosol particles exited the instrument in comparison to breathing, speaking or playing the cross flutes. Furthermore, in agreement with the observation by He et al., the impulse of the exiting cloud was low showing only a small velocity of the cloud ejection. It appears that the trumpet itself reduces the aerosol expulsion by a great amount. Moreover, it remains unclarified to what portion the aerosol remain as particles or condensated liquid inside the trumpet. In a single subject test, a cleaning flow 2 min after the experiment still showed ejection of the remained vapour. In contrast to the trumpets, the absolute aerosol concentrations of the cross flute and the clarinet measured by He et al. [9] was assigned to an intermediate risk level compared with aerosol generation during speaking and breathing. The impulse of the ejected aerosol, however, was largest for the cross flute generating a very high velocity.

The large variability of aerosol expulsion and spreading is attributed to combined effects of different mechanisms of sound production for the three instrument types, mouthpiece and tube structure. Therefore, it seems obvious to transfer the results to instruments of similar construction. It could be assumed that the measurements of the clarinets could be transferred to other reed instruments as the oboe or the cor anglais. Similarly the results of the trumpet could help to estimate the aerosol dispersion of the tuba, the trombone or the horn.

However, in the case of double-reed instruments like the oboe or the cor anglais it seems to be a much tighter seal around the mouthpiece, so that less aerosol may be emitted there. The dispersion from the main outlet can be assumed to be similar, as the straight tube structure is comparable. Regarding the bassoon, it could be assumed that the impulse dispersion is less than that of the oboe due to the curved mouthpiece and the longer tube.

The mouthpiece of the trombone, horn or tuba is similar to that of the trumpet, so that a similarly tight seal with the lips may be ensured and therefore hardly any aerosol would escape. It is conceivable that due to the longer and more tortuous tube system of the above instruments, aerosol particles are deposited and thus less aerosol is emitted by the main outlet. It is worth noting that air turbulence could occur in the trombone due to the trombone slide, which could enlarge the cloud.

The experiment was performed under controlled conditions to decrease external parameters like convectional flow. In order to this spotlights and cameras were positioned in greater distance, the experiment was performed about 5 m to the next wall, the fans of the spotlights were covered and the fan was turned off. However, there was a slight convectional flow mainly directed in positive y-direction. Furthermore, there were single subjects who produced an unexpected large distance during a task, caused by additional convection in the x- and z-direction. The most probable reason for the occurrence of the convection flows is the movements of the subjects just before starting the task. The musicians inhaled the aerosols at the right rear end of the platform with the face heading backwards. After the inhalation, the subjects rotated counterclockwise and stepped at least three steps forward to the musician’s mark. This motion potentially induced a combined rotative and translatory flow which results in a predominate flow in +y-direction, Fig. 2 – 4 b).

After the ejection of the vapor during the task, the aerosols remained in the air, were moved and spread in the room driven by their inertia, decelerated and finally diluted. Three seconds after finishing the tasks the aerosols reached only median distance of .2 m to the front. Thus, the main distance were reached by the initial ejection with large impulse. Consequently, this initial distance has to be considered to define safety distances between instrument players in order to reduce direct person-to-person transmission of potentially infectious particles. However, it could be assumed that aerosol clouds that accumulate over time constitute a high risk for virus transmission [21] if the aerosols are not removed from the air. Hence, beside the definition of safety distances, the aeration rate of rehearsal rooms and concert halls play a critical role for lowering the transmission risk.

The presented study used an artificially added aerosol with a comparable size range of aerosols (250 - 450 nm) for each task. The realistic number of aerosols expelled during playing instruments is, however, much lower and varies among the different instruments. In this respect, it has been found that counting the number of expelled particles with an aerodynamic particle sizer while playing different wind instruments was between 20 and 2400 particles/litre with an average size of 1.9 to 3.1 µm [9]. Thereby the air-jet woodwinds (i.e. cross flute) produced the lowest concentration because a high number of particles were deposited near the inlet [9].

## Limitations

The presented study addresses the aerosol dispersion during time of playing for about 6 to 8 s, so the time for measurement was much too short in comparison to the realistic playing dose in rehearsals or concerts. It could be expected that the accumulation of aerosols in a closed room would be dependent on the playing dose.

The presented study analyzed a short part of the main theme of Ludwig van Beethoven‘s 9^th^ symphony, 4^th^ movement. It cannot be excluded that different melodies or different languages would show different results. However, Schiller’s text with Beethoven’s melody has the great advantage that both, the melody and the text are well known for professional musicians minimizing artifacts due to learning effects.

Owing to the large amount of aerosols, the inhaled gas mixture holds a higher density. This changed some subjects’ self-sensation during playing – presumably due to changes of the resonance properties of the instrument and the gas column. The professionally trained subjects could, nevertheless, compensate these changes during the experiment applying their playing technique also under the experimental circumstances. Moreover, the fact that only professional musicians were included could have influenced the air leakage at the mouthpiece in trumpeters and clarinetists compared to less experienced musicians, because professionals have learned to control air leakage very effectively. However, it can be assumed that the expulsion distances and thus the derived safety distances deviate only slightly from the values measured here.

The number of subjects was too small to perform comparative statistical analyses. It could be hoped that a great number of musicians could be included in future studies in order to test the observed differences and some protective materials to limit the aerosol spread.

## Conclusions

The impulse dispersion of all instruments to the front was found to be greater than in transverse direction to the sides. A distance of 2 m to the front and 1.5 m to the side should be recommended for trumpet and clarinet, 3 m to the front for the cross flutes in an orchestral formation. Our findings could be applied to other orchestral instruments, so other brass instruments such as tuba, trombone or horn may be similar to the trumpet. It could be expected that an oboe and a bassoon have the same or less dispersion than a clarinet. The largest distance occurred during playing the cross flutes. For the risk assessment, the individual playing style, different instrument-specific playing techniques as well as convectional flows in the specific rooms and at least the accumulation of aerosols during playing should also be taken into account.

## Data Availability

The full data set is available from the authors

## Acknowledgements

The authors thank all members of the Bavarian Broadcast for their help in realizing this study.

## Notes

### Competing Interest Statement

The authors have declared no competing interest.

### Funding Statement

The authors have declared no competing interest.

### Author Declarations

LMU Munich etical committe All necessary patient/participant consent has been obtained and the appropriate institutional forms have been archived.

